# The Complex Interplay Between Cancer and the Lung Microbiome

**DOI:** 10.64898/2026.11.04.26367308

**Authors:** Alexander Pohlman, Andrew D Marten, Melline Fontes Noronha, Mark Khemmani, Alan J Wolfe, Zaid M. Abdelsattar

## Abstract

**Background:** The lung fosters a diverse microbiome that may have a complex interplay with underlying diseases, such as lung cancer. However, existing literature has revealed diverse results, likely owing to small sample sizes and diverse sampling methods. In this context, we aim to characterize the lung microbiome and its association with malignancy via multiple sampling methods during lung resection.

**Methods:** Patients undergoing lung resection for confirmed malignancy were prospectively included in the study. During the operation, a bronchial swab and a parenchymal biopsy of lung tissue were collected from the resected specimen and sent for analysis via 16S rRNA gene amplicon (V4) sequencing. An established bioinformatics pipeline was used to determine taxonomic identification and compare these results to clinical and tumor data.

**Results:** There were 83 participants included, with 75 (90.4%) patients having primary lung tumors and 8 (9.6%) having metastatic lesions. Resections included lobectomy (69.9%), wedge (25.3%), and segmentectomy (4.8%). Lung microbiome composition differed by primary malignancy site (lung vs metastatic; p=0.014) and tumor mutations (TP53, p=0.040; KRAS, p=0.054). For primary lung cancer, there was no difference in microbiome composition by tumor histology. A near-significant association was noted for patients with prolonged air leaks and alpha diversity of the lung microbiome (p=0.070).

**Conclusions:** In this study, we demonstrate associations between the lung microbiome and underlying malignancy and tumor characteristics. Validation of these results may assist with diagnostic, prognostic, and therapeutic interventions in the operative and peri-operative management of lung cancer.

## Introduction

Lung cancer is the leading cause of cancer deaths in the United States and surgery remains the gold-standard of treatment.^1,2^ The body’s microbiome has been thoroughly studied in other organ systems, revealing associations with cancer development and surgical outcomes.^3^ In particular, the gut microbiome has been implicated in the incidence of diseases and post-operative complications. For instance, in colon cancer, patients are given decontaminating oral antibiotics prior to resection with the aim of decreasing operative complications.^4^ Yet, minimal literature exists on the interaction between the lung microbiome, lung cancer and curative lung resection.

Studies characterizing the lung microbiome have been limited to small sample sizes, single histologies, or inconsistent sampling techniques and tissue sources.^5,6^ These studies have indicated some important findings, such as a lower biodiversity, and differential microbiomes associated with various tumor-types and characteristics.^5–9^ However, there remains significant variety in results, microbial species identified, and conclusions.^10^ Thus, there is a need for further study of the lung microbiome to determine its association with underlying malignancy and downstream effects on treatment.

In this context, we use two distinct sampling methods to characterize lung airways and parenchyma of patients undergoing lung resection for confirmed malignancy and compare results across patients, tumors, and outcomes. We hypothesize that there would be large diversity in microbiomes with similarities within histology, mutational status, tumor stages, and post-operative complications. The findings provide initial evidence for the role of the lung microbiome on complex disease states.

## Materials and Methods

### Patient Population

Our institutional review board approved this study (IRB#215744, 3/21/22). Adult patients with confirmed primary or metastatic malignancy of the lung undergoing planned surgical resection at our institution were prospectively recruited and consented to the study. Resections included lobectomy, segmentectomy, and wedge resections. All indications for surgery and type of lung resection were determined by the operating surgeon per standard practice. All patients received standard surgical prophylactic antibiotics within one hour of incision time.

### Sampling Technique

We have previously described our microbiome sampling technique.^11^ In brief, both parenchyma (biopsy) and airway (swab) samples were obtained for microbiome analysis. After resection, under sterile conditions, a 1 cm cubic piece of non-cancerous lung parenchyma from the edge of the resected sample was sharply cut and placed in a sterile container for the parenchymal sample. For anatomical resections, the bronchus was sharply cut, and a white-capped Eswab^®^ (Becton Dickinson, Franklin Lakes, NJ) was used to sample the inside of the bronchus and placed into a sterile collection solution. Samples were placed in a biohazard bag, kept at 4°C, and brought directly to the research laboratory, where a nucleic acid preservative (10% AssayAssure, Sierra Molecular, Incline Village, NV) was added to all samples and stored at −80°C. Samples then underwent DNA extraction and sequencing. To assess potential DNA contamination, extraction negative controls (no parenchyma or swab) were processed with the samples.

### Bioinformatic and Statistical Analysis

Adapters were trimmed from the raw sequence reads. Low-quality reads were removed using Cutadapt.^12^ DADA2 software processed the remaining high-quality reads (*i.e.,* filtering, dereplication, chimera removal, construction of an amplicon sequence variant (ASV) abundance table). To achieve taxon identification, BLCA^13^ was used to query the NCBI and Greengenes2 databases.^14^ For each ASV, identification with confidence at the lowest taxonomic level was used for downstream analyses. ASVs with taxa confidence scores less than 69.5% were assigned “unclassified/unknown.” Data is accessible at NCBI BioProject: ID#PRJNA1428797, entitled “Pulmonary Oncogenesis Microbiome Following Surgery.”

*Lysobacter* ASVs were removed as they are known contaminants of the enzyme achromopeptidase used for genome extraction. Low abundance ASVs also were removed. Contaminant ASVs were determined via a scoring method that encompasses results from Decontam,^15^ nonparametric statistical testing (Kruskal-Wallis p≥0.5), mean comparison (*i.e.*, mean counts of samples versus extraction controls), and whether sample read counts were less than 5x greater than controls.

Alpha (within-sample) and Beta (between-sample) diversity indices were computed using statistical packages phyloseq^16^ and vegan^17^ in R (R Foundation for Statistical Computing, Vienna, Austria). Medians with interquartile range (IQR) were presented for alpha diversity measures and within-sample comparisons of differences were performed using Wilcoxon signed rank tests for comparisons between 2 groups or Kruskal-Wallis for comparisons between 3 or more groups. Beta diversity results were compared by abundance-based Bray-Curtis and presence/absence-based Jaccard indices. The analysis included correction for multiple comparisons and statistical significance was determined by Permutational Multivariate Analysis of Variance (PERMANOVA). DESeq2 was used to identify taxa enriched in one variable relative to another ^18^

## Results

### In total, ninety patients were consented and eighty-nine underwent successful resection

Three patients did not have samples taken due to logistical reasons and three patients had benign disease on final pathology. Thus, a total of eighty-three patients were included in this analysis. The study population included 75 (90.4%) primary lung lesions and 8 (9.6%) metastatic lesions. Resection types included 58 (69.9%) lobectomies, 4 (4.8%) segmentectomies, and 21 (25.3%) wedge resections via robotic (85.5%), video-assisted thoracoscopic (12.1%), and open (2.4%) approaches. The most common histologies were adenocarcinoma (60.2%) and squamous cell carcinoma (20.5%; **Table 1)**.

**Table 1.**
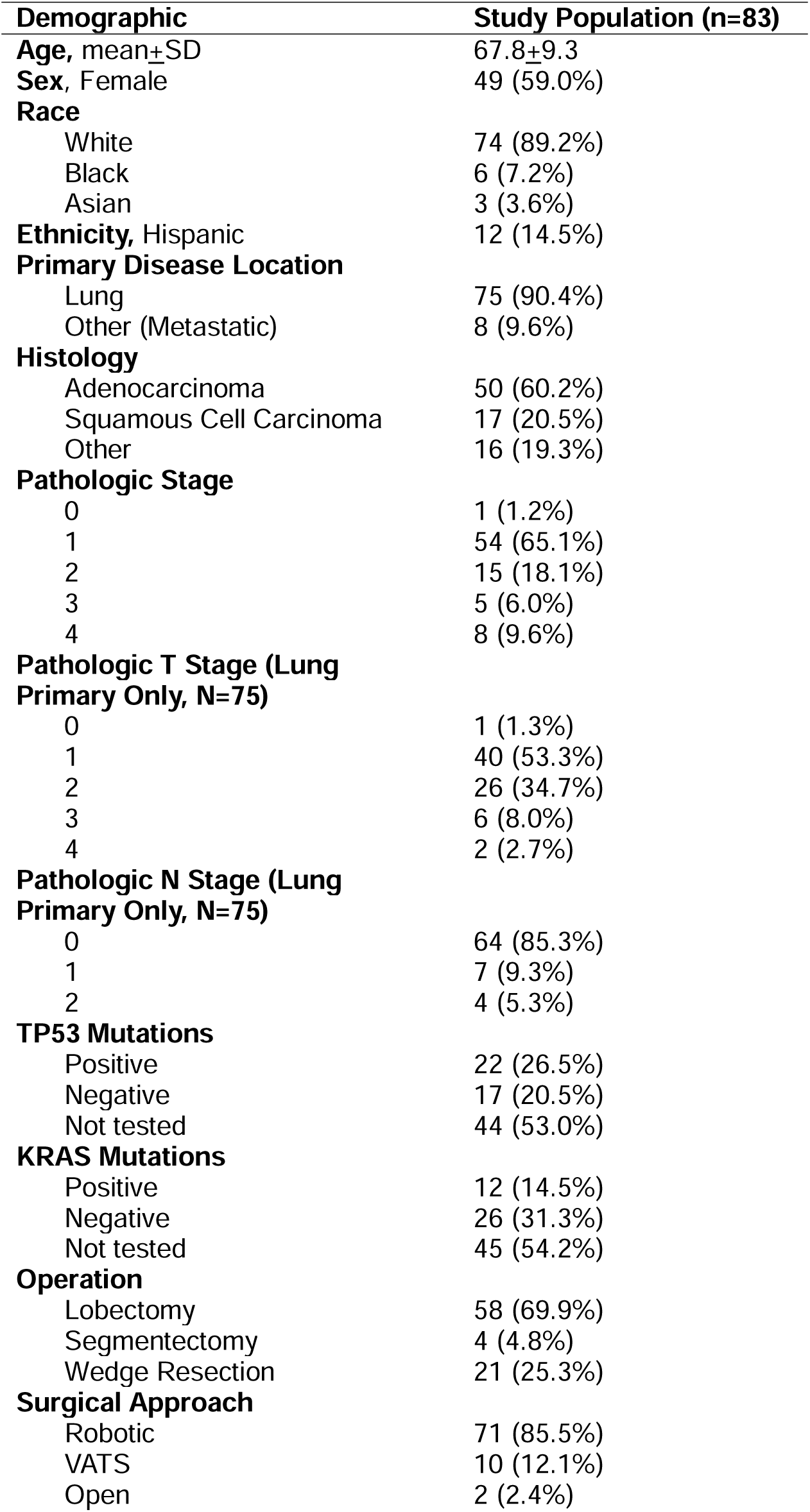
Study population demographics and tumor characteristics.

| **Demographic** | **Study Population (n=83)** |
|---|---|
| **Age,** mean±SD | 67.8±9.3 |
| **Sex**, Female | 49 (59.0%) |
| **Race** |  |
| White | 74 (89.2%) |
| Black | 6 (7.2%) |
| Asian | 3 (3.6%) |
| **Ethnicity,** Hispanic | 12 (14.5%) |
| **Primary Disease Location** |  |
| Lung | 75 (90.4%) |
| Other (Metastatic) | 8 (9.6%) |
| **Histology** |  |
| Adenocarcinoma | 50 (60.2%) |
| Squamous Cell Carcinoma | 17 (20.5%) |
| Other | 16 (19.3%) |
| **Pathologic Stage** |  |
| 0 | 1 (1.2%) |
| 1 | 54 (65.1%) |
| 2 | 15 (18.1%) |
| 3 | 5 (6.0%) |
| 4 | 8 (9.6%) |
| **Pathologic T Stage (Lung Primary Only, N=75)** |  |
| 0 | 1 (1.3%) |
| 1 | 40 (53.3%) |
| 2 | 26 (34.7%) |
| 3 | 6 (8.0%) |
| 4 | 2 (2.7%) |
| **Pathologic N Stage (Lung Primary Only, N=75)** |  |
| 0 | 64 (85.3%) |
| 1 | 7 (9.3%) |
| 2 | 4 (5.3%) |
| **TP53 Mutations** |  |
| Positive | 22 (26.5%) |
| Negative | 17 (20.5%) |
| Not tested | 44 (53.0%) |
| **KRAS Mutations** |  |
| Positive | 12 (14.5%) |
| Negative | 26 (31.3%) |
| Not tested | 45 (54.2%) |
| **Operation** |  |
| Lobectomy | 58 (69.9%) |
| Segmentectomy | 4 (4.8%) |
| Wedge Resection | 21 (25.3%) |
| **Surgical Approach** |  |
| Robotic | 71 (85.5%) |
| VATS | 10 (12.1%) |
| Open | 2 (2.4%) |

We first sorted participants by location of their primary malignancy (lung versus metastatic [Other] tumors) and found that both the biopsy and swab microbiomes differed by beta diversity (biopsy: Jaccard PERMANOVA p=0.0424; swab: Jaccard PERMANOVA p=0.034) (**Figure 1A-B**). For the biopsy samples, DESeq2 analysis revealed 6 ASVs enriched in the metastatic tumors, all members of the phylum Pseudomonadota: ASV35 (*Citrobacter*), ASV38 (*Comamonas*), ASV40 (*Burkholderia*), ASV44 (Hyphomicrobiaceae), and both ASV46 and ASV47 (*Stenotrophomonas*) (**Figure 1C**). For the swab samples, DESeq2 analysis revealed 7 ASVs enriched in metastatic disease, 5 members of the phylum Pseudomondota [(*i.e.,* ASV31 (*Eikenella*), ASV38 (*Comamonas*), ASV52 (*Pseudomonas*), ASV54 (*Symbiopectobacterium*), ASV72 (*Ralstonia*)], 1 member of Actinomycetota [ASV11 (*Tropheryma*)], and 1 member of Cyanobacteriota [ASV39 (*Obscuribacter*)] (**Figure 1D**). Lung versus metastatic disease samples did not differ by alpha diversity.

**Figure 1.**
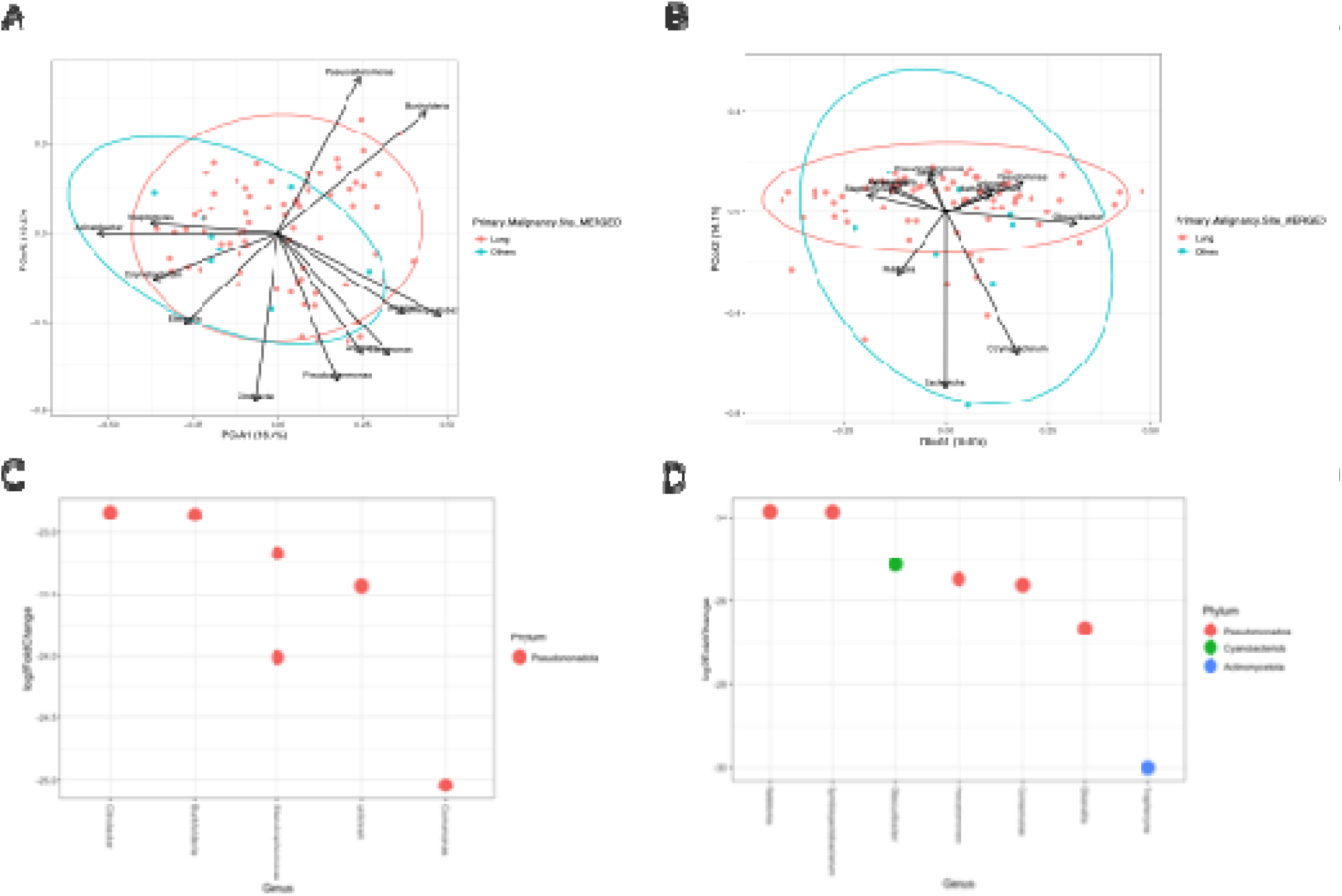
Comparison of microbial diversity in samples by location of the primary malignancy (lung vs others [malignant]). (A & B) Biplot of Principal Coordinates Analysis (PCoA) scores based on the Jaccard index, revealing significant differences in beta diversity for both the biopsy (PERMANOVA p=0.0424) and swab (PERMANOVA p=0.034) samples, respectively. (C & D) DESeq2 Analysis demonstrating enriched species in the primary malignancy site for the biopsy and swab samples, respectively. Positive Log_2_FoldChange enriched in other primary malignancy sites (metastatic); negative Log_2_FoldChange enriched in lung primary tumors

We then sought to measure how oncogenes and other cancer-related clinical variables may be associated with microbiome composition. First, we assessed microbial diversity with respect to the presence (POS) or absence (NEG) of TP53 mutations, a vital tumor suppressor gene. Based on the Bray-Curtis dissimilarity index, there was a significant difference in beta diversity (p=0.040) between POS/NEG TP53 in swab samples (**Figure 2A**). However, we did not detect a significant difference in alpha or beta diversity in biopsy samples. DESeq2 revealed 8 ASVs differentially enriched between POS/NEG TP53 swab samples, all members of the phylum *Pseudomonadota* (**Figure 2B**). ASV52 (*Pseudomonas*), ASV54 (*Symbiopectobacterium*), and ASV59 (*Methylobacterium*) were enriched in TP53 wild-type samples. In contrast, ASV38 (*Comamonas*), ASV47 (*Stenotrophomonas*), ASV72 (*Ralstonia*), and ASV26/ASV33 (*Pseudomonas*) were enriched in TP53-mutant samples. We also measured significant differences in alpha-diversity between patients with and without KRAS mutations in biopsy samples (**Figure 2C**).

**Figure 2.**
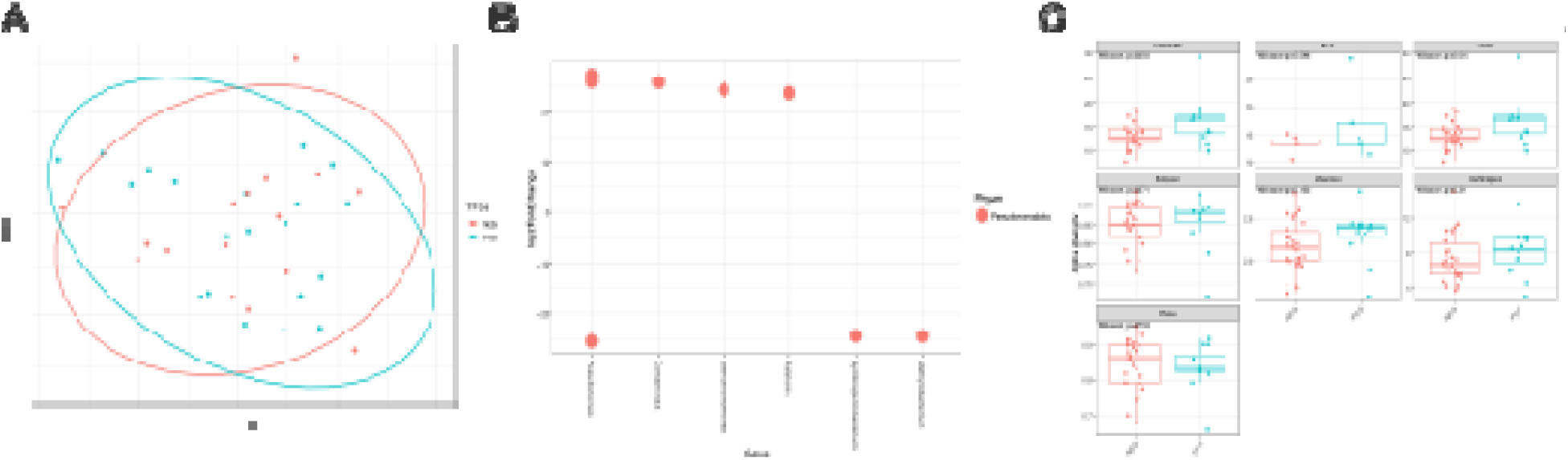
Comparison of microbial diversity in swab samples dichotomized by the status of the p53 gene (TP53): positive (POS) versus negative (NEG). (A) Principal Coordinates Analysis (PCoA) based on the Bray-Curtis dissimilarity index, revealing significant differences in beta diversity (PERMANOVA p=0.0398). (B) DESeq2 Analysis revealing enriched species in those with and without TP53 mutations. Positive Log_2_FoldChange enriched in positive (TP53-mutants); negative Log_2_FoldChange enriched in negative (TP53 wild-type). (C) Alpha diversity of tumors with (POS) and without (NEG) KRAS mutations, revealing increased richness in those with mutations (Observed and Chao1, p=0.054).

We evaluated several other clinical variables to gain a better understanding of how lung cancer may influence the microbiome or vice versa. When comparing the two most common lung histologies (adenocarcinoma and squamous cell carcinoma) we found no significant difference in alpha diversity (all p>0.05, **Supp. Figure 1**). We also saw no significant difference by tumor T stage in lung primary tumors: T0/T1 vs T2 vs T3/T4 (**Supp. Figure 2**) and N stage: node negative (N0) versus node positive (N1/N2) disease (**Supp. Figure 3**). Similarly, there was no difference in beta diversity for lung histology, T stage or N stage.

**Figure 3.**
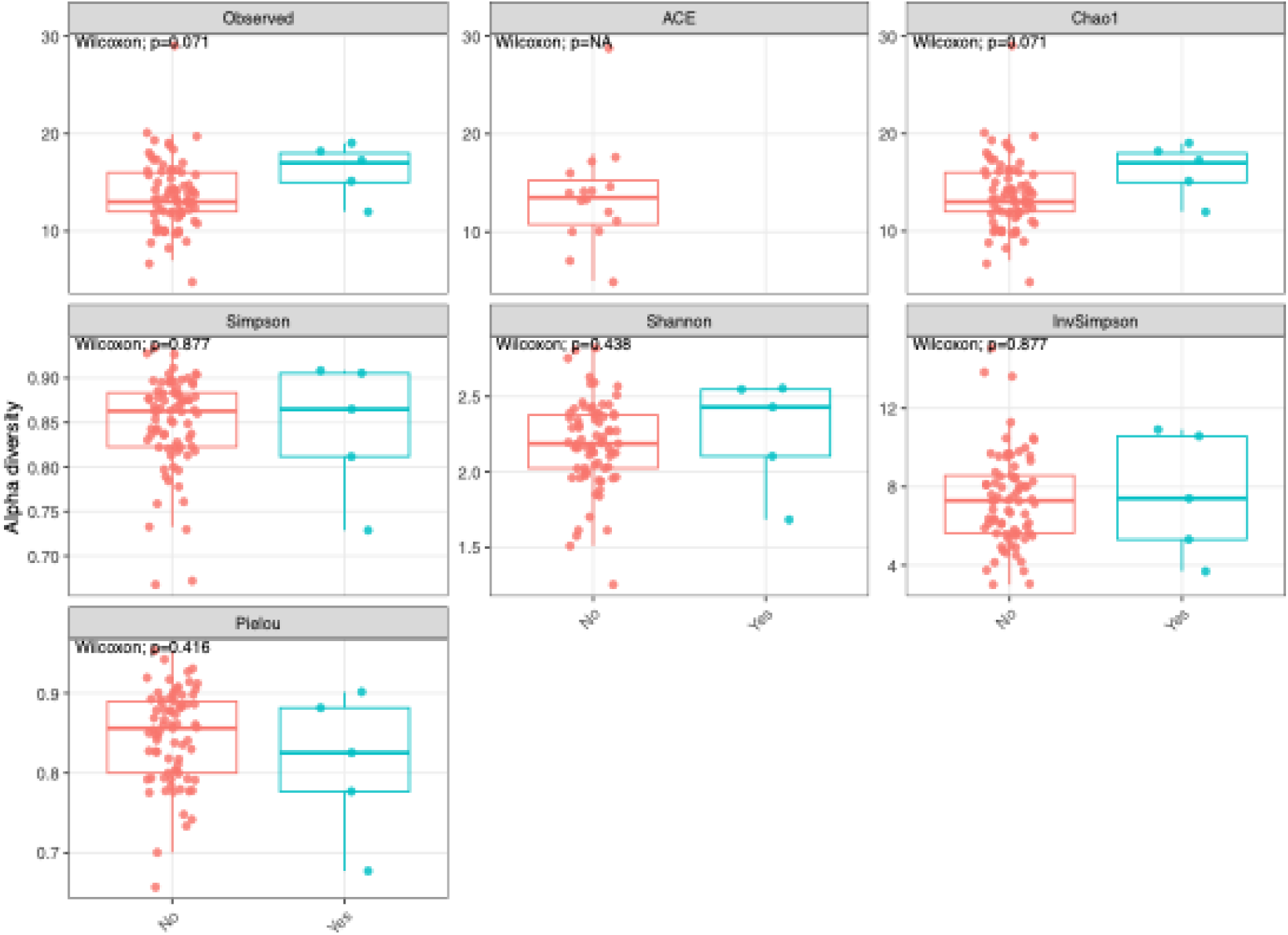
Comparison of microbial alpha diversity for patients that did and did not develop a post-operative prolonged air leak (PAL > 5 days) showing near significant differences in measures of richness (Observed and Chao1, p=0.071).

We also quantified whether differences in the microbiome correlated with any definitive clinical outcomes. First, we analyzed pulmonary complications, including pneumonia and prolonged air leaks. Our analysis of post-operative pneumonia revealed a significant difference in beta diversity (Bray-Curtis p=0.026); however, there were only three cases of pneumonia, which limited this analysis. In a comparison of patients with prolonged air leaks (defined as air leaks persisting beyond 5 days), there was a trend toward significance for alpha diversity (richness, p=0.071; **Figure 3**). However, there were no differences seen in composite measures of cardiovascular complications, including myocardial infarction, hemothorax, or atrial fibrillation (**Supp. Figure 4**; all p>0.05) and overall complications, which was the composite of any pulmonary or cardiovascular complication (**Supp. Figure 5**).

## Discussion

In this prospective observational study of the lung microbiome in patients undergoing resection for primary or metastatic malignancy of the lung, we found that the lung microbiome surrounding tumors (1) differs by primary cancer disease site (i.e., lung versus metastatic disease); (2) varies with tumor mutations; and (3) may have a complex interplay with surgical factors and outcomes. Ultimately, there is a complex interplay between the lung microbiome and malignancy that may impact oncologic and surgical outcomes. A better understanding of these interactions could improve risk-stratification and treatment of tumors involving the lung.

There is a growing body of evidence surrounding the association between the lung microbiome, cancer development and progression. In this study, we found that primary and metastatic disease were associated with different microbiome compositions. There are several possible explanations for this. For one, prior studies have shown that some bacteria are carried within tumor cells,^19^ so it’s possible that metastatic tumors carried bacteria with them from other organs, as has been shown with *Fusobacterium*.^20^ These bacteria may not only be incidentally carried with these tumors, but there may be a mechanism by which they promote the spread of disease. Prior studies have shown that intracellular microbiota can promote metastatic colonization,^21^ or facilitate the epithelial-to-mesenchymal transition whereby tumor cells lose their adhesion to neighboring cells.^22^ In this study, we found enrichment of *Stenotrophomonas* in metastatic tumors. Prior studies have shown increased rates of *Stenotrophomonas* surrounding metastatic colorectal tumors to the lung and have linked this microbe with an overall poor prognosis and cancer progression.^23^ Some prior studies have linked toxins produced by certain bacterial species, such as *Citrobacter rodentium* and *Campylobacter jejuni* to DNA damage and carcinogenesis.^24^ We similarly, found *Citrobacter* to be enriched around metastatic tumors, which further supports this hypothesis. We also found enrichment of *Ralstonia* and *Comamonas* in patients with metastatic disease, which have previously been demonstrated in biliary tract cancers.^25^ Thus, our results support findings in other disease sites and studies, while expanding upon them.

However, it’s also important to note that the cause-effect relationship between these tumors and the microbiome is unclear. It’s possible that tumor cells may produce metabolites or microenvironmental factors that are more conducive to specialized bacterial growth, thus favoring certain species.^26^ On the other hand, the microbiome itself may foster a microenvironment that favors the development of primary lung cancer or metastatic lesions. In this prospective, observational study, it is difficult to determine causation but, regardless, these associations may serve as important diagnostic, prognostic, and therapeutic markers.^27^ Further research may help elucidate these findings.

There were also several important relationships between the microbiome and mutations within these tumors, including TP53 and KRAS. Similar differences were seen with TP53 mutations in prior studies, but one of these prior studies only found this difference in adenocarcinoma and the other only found it in squamous cell carcinoma,^8,28^ so our results confirm and expand on these prior findings. However, when identifying specific bacterial species, there were some differences in our results compared to prior studies. For example, we found that *Ralstonia* was enriched in TP53 wild-type (negative) samples whereas one prior study found enrichment of *Ralstonia* in TP53-mutant (positive) samples.^28^ Other studies have identified enrichment of *Ralstonia* in patients with lung adenocarcinoma,^29^ so it appears that this genus has an association with cancer but it may have a more complex interplay with certain histologic and mutant subtypes.

Similarly, our study revealed associations with *KRAS* mutations, which were previously only demonstrated in the gut microbiota of patients with colorectal cancer.^30^ Thus, our study now expands this finding to an entirely new disease site. There were some differences in our findings as well; for example, prior studies found differences in beta diversity by KRAS status, including the enrichment of *Faecalibacterium* in *KRAS*-mutants versus enrichment of *Bifidobacterium* in *KRAS* wild-type tumors.^31^ Meanwhile, we found differences in alpha diversity, rather than beta diversity. Thus, it seems that differences exist, but the significance and specific bacterial species and type of diversity may vary by tumor and underlying mutations. Given that these associations have now been found in multiple cancer types and mutations, it’s unlikely that they occurred by chance. Rather, it’s likely that there are clinically meaningful interactions between tumor mutations and the surrounding microbiome that may represent both prognostic and therapeutic targets for cancer treatment. In fact, prior studies have shown that some of these mutations can have direct impacts on neoadjuvant and adjuvant treatments, such as immunotherapy. For example, TP53 and the microbiome have both been shown to predict response to immunotherapy,^28,32–36^ so isolating bacteria that may impact neoadjuvant or adjuvant treatment efficacy could have significant implications on clinical practice. Some studies have suggested that certain bacteria may promote pro- or anti-tumoral environments via inflammatory, immunomodulatory, or genetic mechanisms, which could ultimately impact therapeutic targets.^37–41^ If these microbes directly impact the efficacy of treatment, one could attempt to eradicate them with targeted antimicrobial medication or elect to treat the patient with a different therapy option.

We also identified several relevant null findings, including T/N stages and histology. Existing literature has shown an association between advanced disease and differences in the lung microbiome,^42–44^ so it’s feasible that this null finding was due to sampling distance from the tumor as we did not take tissue directly from tumors but rather took surrounding lung parenchyma and airway samples from the edges of resected specimens. Overall, it seems that much more variety exists in the lung microbiome of individuals with lung primaries than with metastatic disease, so future studies directly comparing specific tumor types and tumor samples to surrounding parenchyma at different distances could help elucidate these findings within lung primary tumors.

There were a few notable findings in the lung microbiome of patients experiencing peri-operative complications. Until this time, the study of the human microbiome and surgical outcomes has primarily focused on the gut microbiome and colorectal cancer, showing that a healthy microbiome may improve recovery times, bowel function, and decrease surgical site infections.^45^ However in the lung, the microbiome could similarly impact recovery. In the case of lung resections, there is a very similar staple line across an epithelial surface that requires proper wound healing. If these cut surfaces do not heal appropriately, prolonged air leaks may ensue. We found a near-significant difference in the microbiome of patients experiencing prolonged air leaks. It’s likely that a larger study population may be needed to confirm this finding, but there may be a direct mechanism where this may occur. For example, collagenase producing bacteria, such as *Pseudomonas*, which was enriched in several of our samples, could lead to increased collagen breakdown, poor wound healing, and leaks.^46^ Thus, these bacteria could serve as a target for selective decontamination prior to an operation in an attempt to minimize this complication.

Additionally, there were three patients who developed post-operative pneumonia in our study. These samples demonstrated an altered microbiome compared to the rest of the cohort, but the small number of patients experiencing this complication make it difficult to reliably analyze in the current study. Prior studies have implicated certain oral flora, such as *Fusobacterium* and *Enterococcus*, in the likelihood of developing a post-operative pneumonia.^47^ Although we could not directly identify which species were enriched in patients with post-operative pneumonia in our study, several of the taxa identified in our other analyses have been found in common pulmonary conditions, such as *P. aeruginosa, Acinetobacter*, *Burkholderia, Comamonas, Ralstonia* and *Stenotrophomonas* in cystic fibrosis, and *P. aeruginosa* in nosocomial infections.^48–51^ *Stenotrophomonas maltophilia* is also thought to be a nosocomial pathogen associated with pneumonia and has been reported to be a marker of lung disease severity.^52,53^ If future studies with larger cohorts confirm a difference in the lung microbiome associated with post-operative pneumonia, it would further support the use of pre-operative administration of antimicrobials for selective decontamination prior to lung resection.

This study has several limitations. First, our patient sample was limited to a single institution, which limits generalizability. However, this also improved our ability to compare patients given similar demographics, geographic locations, and potential environmental exposures. Second, we only saw trends in certain outcome measures, such as post-operative air leaks, so it’s possible that with a larger study cohort these differences could become clearer. Third, patients received standard surgical antimicrobial prophylaxis within one hour of their operations. Although these antibiotics could kill susceptible microbes, most surgical prophylactic antibiotics, such as cefazolin, kill bacterial cells by blocking cell wall synthesis rather than causing cell rupture and nucleic acid damage so their DNA is typically still detected by 16S rRNA gene sequencing.^54^ Finally, we aimed to minimize the risk of contamination through the use of negative controls and bioinformatic decontamination; however, some of the genera identified are known members of the kitome (i.e., contaminants associated with sample processing),^55^ so we interpret our findings cautiously and, in general, our findings, discussion, and conclusions focused on genera that have been reported as lung-associated. Additionally, since the noted organisms were amongst the most relatively abundant in our analyses, they are unlikely to be contaminants. Whether any of these particular genera are signatures of cancer-specific factors will require larger and confirmatory studies.

## Conclusions

In conclusion, there is a complex interplay between the lung microbiome and underlying malignancy. There are several important associations with the microbiome and cancer-specific factors, including primary versus metastatic disease and tumor mutations that could serve as important diagnostic, prognostic, and therapeutic targets for future medical and surgical management of lung cancer. Further validation of the lung microbiome and surgical outcomes could provide evidence for the selective decontamination of the lung via inhaled antibiotics prior to operations.

## Data Availability

Data is accessible at NCBI BioProject: ID#PRJNA1428797, entitled Pulmonary Oncogenesis Microbiome Following Surgery.

## Acknowledgements

We would like to thank the surgeons who collected specimens, including Drs. Julia Coughlin, James Lubawksi, Wissam Raad, and Wickii Vigneswaran. We would also like to thank clinical/research personnel who assisted with sample collection and processing, including Raymond Verm, Madison Lozanoski, Mohamed Gadelkarim, Ayham Odeh, Bilal Odeh, Joseph Zywiciel Jr, Omar Zahra, and Germaine Harvey. Finally, we are deeply grateful to all the patients whose participation made this study possible.

## Author Contributions (CRediT)

**Alexander Pohlman**: methodology, validation, investigation, data curation, visualization, writing – original draft, writing – review and editing. **Andrew Marten**: methodology, validation, data curation, writing – original draft, writing – review and editing. **Melline Fontes Noronha**: Methodology, software, validation, formal analysis, data curation, writing – original draft, writing – review and editing, visualization. **Mark Khemmani**: Methodology, validation, investigation, writing – original draft, writing – review and editing. **Alan Wolfe**: Conceptualization, methodology, validation, resources, project administration, supervision, writing – original draft, writing – reviews and editing, funding acquisition. **Zaid Abdelsattar**: Conceptualization, methodology, validation, resources, project administration, supervision, writing – original draft, writing – reviews and editing, funding acquisition.

