## Supplemental Figures 1-5 for "The Complex Interplay Between Cancer and the Lung Microbiome": Cancer and Lung Microbiome Supplemental Material.docx

**Supplemental Figure 1.** Alpha diversity analysis for lung histology: adenocarcinoma versus squamous cell carcinoma, revealing no significant difference in richness, evenness, or overall composite measures: (A) Biopsy samples; (B) Swab samples

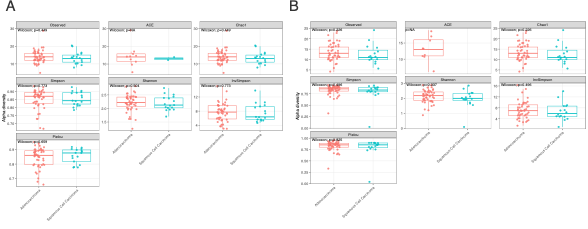

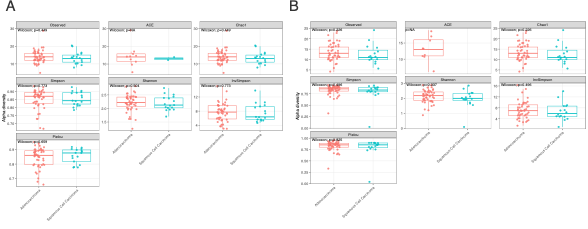

**Supplemental Figure 2.** Alpha diversity analysis for comparing Tumor T stages: 1 (T0-T1), 2 (T2), and 3 (T3-T4). No significant difference was found in richness, evenness, or overall composite measures of alpha diversity: (A) Biopsy samples; (B) Swab samples

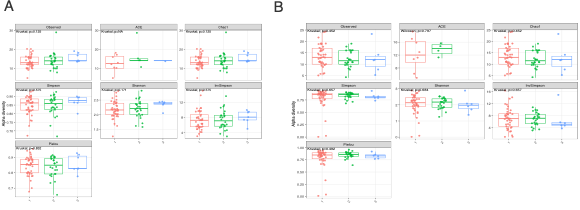

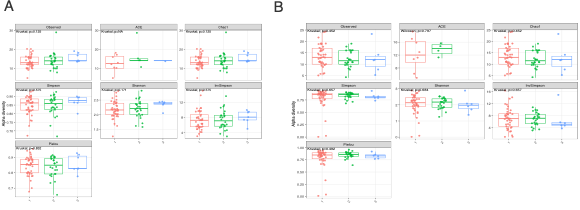

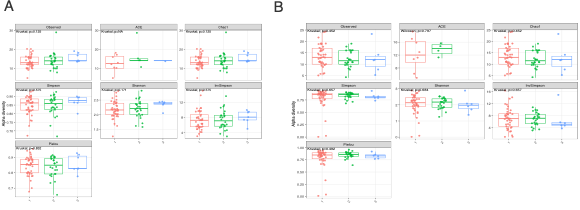

**Supplemental Figure 3.** Alpha diversity analysis for Nodal (N) stage: 0 (N0), 1 (N1+). No significant difference was found in richness, evenness, or overall composite measures of alpha diversity: (A) Biopsy samples; (B) Swab samples

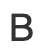

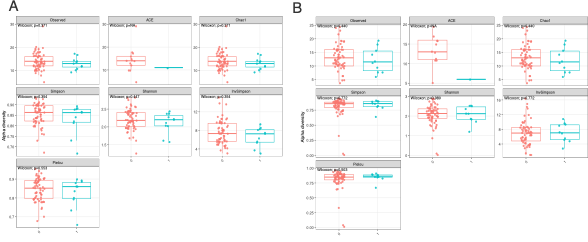

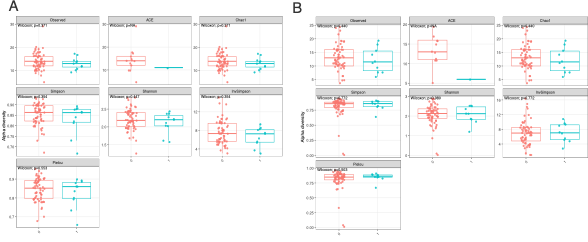

**Supplemental Figure 4.** Alpha diversity analysis comparing those with and without any post-operative cardiovascular complication: 0 (no cardiovascular complication) and 1 (any post-operative cardiovascular complication). Cardiovascular complications included atrial fibrillation, myocardial infarction, and hemothorax. No significant difference was found in richness, evenness, or overall composite measures of alpha diversity: (A) Biopsy samples; (B) Swab samples

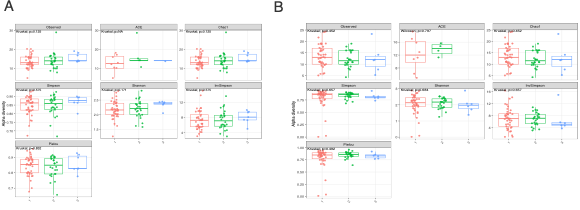

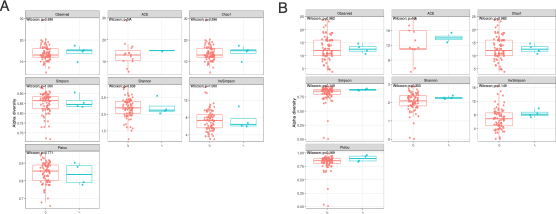

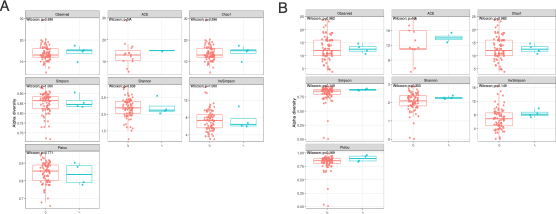

**Supplemental Figure 5.** Alpha diversity analysis comparing those with and without any post-operative complication: 0 (no complications) and 1 (any post-operative complication). No significant difference was found in richness, evenness, or overall composite measures of alpha diversity: (A) Biopsy samples; (B) Swab samples

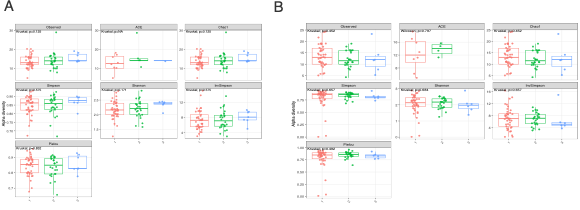

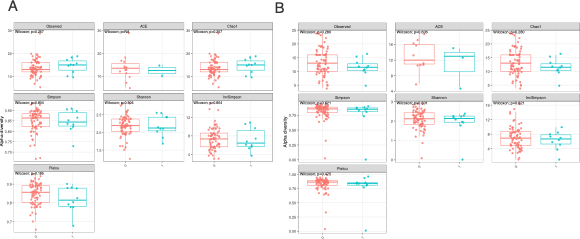

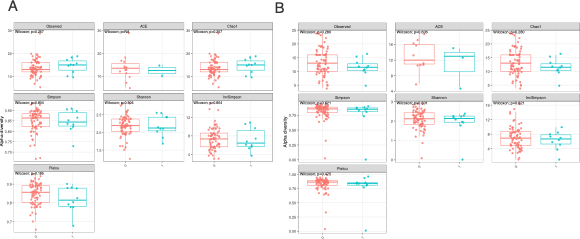
